# Prevalence and factors associated with hepatitis B and C virus infections among female Sex workers in Ethiopia: Results of the national biobehavioral Survey, 2020

**DOI:** 10.1101/2022.05.24.22275494

**Authors:** Bira Bejiga Bedassa, Gemechu Gudeta Ebo, Jemal Ayalew, Jaleta Bulti Tura, Feyiso Bati Wariso, Sileshi Lulseged, Getachew Tollera, Tsigereda Kifle, Saro Abdella

**Affiliations:** HIV/AIDS disease research team, TB and HIV/AIDS Disease Research Directorate, Ethiopian Public Health Institute, Addis Ababa, Ethiopia; TB disease research team, TB and HIV/AIDS Disease Research Directorate, Ethiopian Public Health Institute, Addis Ababa, Ethiopia; Faculty of Medicine, College of Health Sciences, Addis Ababa University, Ethiopia; Deputy Director General, Ethiopian Public Health Institute, Addis Ababa, Ethiopia; Director General, Ethiopian Public Health Institute, Addis Ababa, Ethiopia

**Author notes:** These authors contributed equally to this work (Bira Bejiga Bedassa, Gemechu Gudeta Ebo, Jemal Ayalew, Saro Abdella). This author contributed less but substantially (Jaleta Bulti Tura, Feyiso Bati Wariso, Sileshi Lulseged, Getachew Tollera, Tsigereda Kifle).

**Keywords:** hepatitis B, hepatitis C, female sex workers, syphilis, Ethiopia

## Abstract

**Background:** Hepatitis B and C virus infections are endemic diseases in sub-Saharan Africa, the region with the highest prevalence of these infections in the world. Female sex workers are exposed to sexually transmitted infections, including hepatitis B and C, because of their high-risk sexual behavior and limited access to health services. There is no data on national prevalence estimate on hepatitis B and C virus infections among female sex workers in Ethiopia, a critical gap in information this study aimed to fill.

**Methods:** This was a cross-sectional, biobehavioral, national study conducted from December 2019 - April 2020 among 6085 commercial female sex workers aged ≥15 years and residing in sixteen (n=16) regional capital cities and selected towns of Ethiopia. Blood samples were collected for hepatitis B and C virus serological testing from the participants. The data were collected using an open data kits (ODK) software and imported into STATA version16 for analysis. Descriptive statistics (frequencies and proportions) were used to summarize data on the study variables. Bivariate and multivariate logistic regression analyses were conducted to determine the strength of association between independent variables (risk factors) and the outcome (hepatitis B and C virus infection). Adjusted Odd ratio (AOR) was used to determine independent associations, 95% confidence interval to assess precision of the estimates, and a *P* value ≤ 0.05 was considered statistically significant.

**Results:** The prevalence of hepatitis B and C infections among the 6085 female sex workers was 2.6% [(95% CI (2.2,2.8)] and 0.5% [(95% CI (0.4,0.7)], respectively. Female sex workers who had 61-90 and ≥91 paying clients in the past six months [(AOR=1.66; 95% CI, (0.99, 2.79); *P*=0.054] and [(AOR=1.66 95% CI, (1.11, 2.49); P=0.013], respectively, age at first sex selling of 20-24 and >25 years [(AOR=1.67; 95% CI, (1.14, 2.44); *P*=0.009)] and [(AOR=1.56; 95% CI (1.004, 2.43); *P*=0.048)], respectively, known HIV positive status [(AOR=1.64; 95% CI (1.03, 2.62); *P*=0.036] were significantly associated with the prevalence of hepatitis B virus infection. Similarly, hepatitis C was significantly associated with, age at first sex ≤15 years and age 16-20 years [(AOR=0.21; 95%CI (0.07,0.61); *P*=0.005)] and [(AOR=0.18; 95% CI (0.061, 0.53); *P*=0.002)], respectively, known HIV positive status [(AOR=2.85; 95%CI (1.10,7.37); *P*=0.031)] and testing positive for syphilis [(AOR=4.38; 95% CI (1.73,11.11); *P*=0.002)], respectively.

**Conclusion:** This analysis reveals an intermediate prevalence of hepatitis B and a low prevalence of hepatitis C infection among female sex workers in Ethiopia. It also suggests that population groups with like female sex workers are highly vulnerable to hepatitis B, hepatitis C, and other sexually transmitted infections. There is a need for strengthening treatment and prevention interventions, including immunization services.

## INTRODUCTION

Viral hepatitis is an important public health problem globally, and hepatitis B virus (HBV) and hepatitis C virus (HCV), in particular, are endemic in developing countries (1). It is estimated that 325 million people were living with HBV and HCV in 2019 globally (2). In 2013, viral hepatitis was the seventh-highest cause of mortality in the world and it was responsible for an estimated 1.4 million deaths per year, mostly from hepatitis-related liver cancer and cirrhosis; approximately 47% of the deaths were attributable to HBV, 48% to HVC, and the remainder to hepatitis A and E infections (3). According to the 2017 Global Hepatitis Report, viral hepatitis in general caused 1.34 million deaths in 2015, a figure that was higher than the deaths caused by HIV infection and comparable with that caused by tuberculosis (4).

In the African Region, HBV infection is highly endemic and affects an estimated 5%-8% of the population, mainly in West and Central Africa (5). People living with HIV (PLHIV) are at high risk of becoming ill and dying from hepatitis. Some 2.6 million PLHIV are co-infected with HBV, and some 2.3 million with HCV (6). The WHO 2019 Progress Report on HIV, viral hepatitis, and sexually transmitted infections (STI), showed, in the sub-Saharan Africa (SSA), there were over 60 million cases of chronic hepatitis B and over 10 million cases of chronic hepatitis C infections (7).

In SSA, female sex workers (FSW) had a high-risk behavior and remain important in terms of transmission of HBV, HCV, HIV/AIDS and other STI acquisition and transmission. This could be because FSW have numerous sex partners and they were engaged in unprotected and other forms of sex that cause contact with body fluids of a partner who has STI. FSW are often in a weaker position to negotiate safe sex because of social, economic, cultural and legal reasons (8). Commercial sex work (CSW) is a high-risk activity associated with HBV, HCV, and several other STI (6, 9). The higher risk of getting infected with HIV and other STI, such as syphilis and hepatitis among FSW is primarily associated with the high number of sexual partners and increased frequency of unprotected sex. Several studies have shown that low adherence to condom use, multiple sexual partners, unsafe sexual practices, illicit drug use, and co-infection with other STI increase the risk of HBV and HCV transmission. FSW also have a higher risk of contracting STI from their non-paying partners than from their paying clients (10,11).

In Ethiopia, FSW carry a disproportionate burden of HBV, HCV, and HIV infection. According to the Ethiopian Demographic and Health Survey (EDHS) 2016 report, the marked regional variation that was driven by most at-risk populations (MARPS) indicates that urban areas and females are more affected than rural areas and males, respectively (12). Small towns are also becoming hotspots and can potentially bridge further the spread of the HIV and HBV infections to rural settings, where the female are twice more affected than males (12).

Ethiopia is in the region where HBV prevalence is considered hyper-endemic with a prevalence of between 8%-12%, and that of HCV prevalence is estimated at not less than 2.5% (13). An earlier study conducted in Ethiopia reported that 12% of hospital admissions and 31% of the mortality on the medical wards in Ethiopian hospitals were due to chronic liver disease (CLD) (13). However, there isn’t much done and the available data on associated chronic liver disease or hepatocellular carcinoma are not sufficient.

There are limited data from isolated studies showing the prevalence of HBV in Ethiopia - Hawassa 9.2% (14), Gonder 28.9% (15), Mekelle 6% (16), Dessie 13.1% (11), and northwest Ethiopia (11.9%) (17). FSW have been identified as a population group with the highest risk for STI, including HBV and HCV, and should perceive priority in the national HIV/AIDS program (18). Although there is an ongoing HBV, HCV, and other STI program in the country, there is no national data among FSW to determined HBV and HCV prevalence and driving factors. Therefore, the current study was conducted to explore the prevalence of HBV and HCV infections and identify the factors associated with these infections among FSW in Ethiopia.

## MATERIALS AND METHODS

### Study setting and population

The study was done in Ethiopia, a country divided into eleven regions and two city administrations, and with a population of 119.8 million, some 80 ethnic groups, and a land area of 1.0 square kilometers, and an average population density of 121 inhabitants per square kilometers (19). It had a gross domestic product growth of 6.1% in 2020 and a per capita income of USD 850 (20). The median age of the population is 19.6 years, population growth rate 2.56%, total fertility rate (TFR) 4.6, infant mortality rate (IMR) 41per 1,000 live births, and adult literacy rate of 49% in 2016 in 2016, when the country started a national viral hepatitis prevention and control program (21). Regional capital cities and selected towns with the highest number FSW (hotspots), including Adama, Addis Ababa, Arba Minch, Bahir Dar, Combolcha/Dessie, Dilla, Dire Dawa, Gambella, Gonder, Harar, Hawassa, Jimma, Logia/Semera, Mizan, Nekemite, and Shashemane were involved in the study.

### Study design and period

This was a crass-sectional, nation-wide, biobehavioral study conducted among FSW aged ≥15 years during the period from December 2019 - April 2020.

### Target population

The target population of the study is all FSW living in cities and towns in Ethiopia, and the sampling frame is the list of FSW residing in the regional capitals and selected towns with FSW-hotspots in Ethiopia.

### Study population

FSW aged ≥15 years residing in regional capitals and selected towns with FSW-hotspot or who worked in these cities and towns in the last one month preceding the survey. The survey included both fixed (venue-based) and floating (street-based) FSW.

### Inclusion and exclusion criteria

We included women aged ≥15 years, who received money/other benefits in exchange for sex with four or more people within the last 30 days, agree to participate in the survey including interviewing and biological testing, able to provide informed consent and communicate in one of the survey languages, had a valid coupon provided by the study team, and residing or working in the survey city or town for the last one month.

### Sample size and sampling procedure

The sample size was determined by single population proportion formula

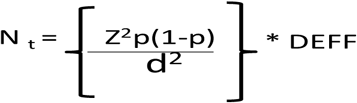

using 95% confidence interval, α = 0.05, margin of error of 35% (d), and proportion (p) (22) of 2%, and DEFF = design effect with a replacement for non-responders. With these assumptions, the minimum desired sample size of FSW in sixteen (n=16) major regional capitals and selected towns with FSW hotspots was 6085 after adding 10% contingency. This was divided and assigned to 16 sites proportionate to population size.

Specific hotspot areas for FSW were identified during tools and procedures pretesting with support from HIV/AIDS Prevention and Control office (HAPCO), woreda (district) health offices, and drop-in-clinics (DICs), local organizations working with FSW. We used a respondent-driven consecutive sampling using a standardized questionnaire for recruitment of study participants. The local organizations assisted in identifying the initial respondents of the survey, referred to as “seeds”. The number of seeds for each site was determined based on the result of a formative assessment. Five “seeds” for each site with allocated sample of <450, six-eight seeds for each site with sample 450-900, and 12 seeds for each site with sample of 1101 were recruited. The “seeds” were selected based on the type of sex worker, age category, and geographic location of the site. These include those FSW who were bar- and/or hotel-based, red lighthouses, local drinking houses, street-based and hidden (cell phone-based).

FSW with a known social network were given each three coupons for use to invite her friends or other FSW contacts who were in her network. This approach helped in reaching as many eligible FSW as possible. The coupon remained active from the day it was given to the potential participant and expired after two weeks or if the study was completed earlier. We used anonymous fingerprint-based code obtained using biometric fingerprint scanners to ensure that all respondents participated only once. This was not linked to the biobehavioral questionnaire and was used only for avoiding multiple enrollments.

Damaged, mutilated, not readable, photocopied, not sealed/stamped coupon was considered not valid. Each participant coming to the study site would need to bring her coupon that was identified by specific number given by the referring person. New participants were given coupons and asked to recruit three additional acquaintances. This process continued until the desired sample size was achieved and respondent-driven sampling (RDS) equilibrium condition attained. Progress towards reaching the equilibrium was monitored by using key parameters including current HIV status, type of sex work, and consistent condom use.

### Data collection procedure and data quality management

Data were collected using a pre-tested structured questionnaire initially developed in English and then translated into the local language (Amharic) was entered onto open data kits (ODK) software. Training was provided to the study team, coordinators, interviewers, blood sample collectors, coupon managers (for RDS), receptionists (for RDS) and accompanying referral liaisons. The training included different topics with a focus on the survey sampling methodology, procedures, and data collection tools, and overall study site management. Data collection tools and questionnaires were pretested in a pilot survey in Bishoftu town, a site not included in the main study. Feedback from the pilot was used to finalize the data collection tools, capturing process, field logistics, and operational procedures. The survey used whole blood for the rapid HIV, hepatitis-B and syphilis testing. After collecting whole blood of 5 ml using EDTA tube; HIV testing, Hepatitis B surface antigen (HBsAg), hepatitis C antibody (HCVAb) and syphilis testing were performed right after sample collection. Then after centrifuging the whole blood, the plasma was separated and a liquated in two a 1.8 ml preprinted labeled nunc tube for viral load quantification and quality control testing.

### Biological analysis

HBsAg and HCVAb were screened by using a rapid test kit according to manufacturer principles and procedure. Testers were trained on how to use the test kit and control lines. All invalid results were repeated. Quality control (QC) panels consisting of a positive and negative control specimen were done in parallel with the testing procedure to ensure test kits were performing correctly. Other STI, including HIV and syphilis, were tested using the national rapid testing algorithm and the results were returned during the second study visit. Syphilis was screened using Chembio Dual Path Platform (DPP) Syphilis Screen and Confirm Assay, according to manufacturer principles and procedures in the kit insert.

### Data Analysis

The data was collected using the ODK software on tablet computers, and was exported to MS-EXCEL, cleaned, and imported to STATA Verssion16 for analysis. The RDS recruitment process (Tree of recruitment), assessment of the RDS assumptions, and RDS weight generating were implemented using the RDS package inbuilt in R statistical software (23). Homophily and convergence, the common assumptions in RDS, were checked in HIV status, consistent condom use, and type of FSW and met the RDS criteria. The RDS weights were exported using the RDS-II function to STATA and merged with the whole dataset for further analysis. Descriptive statistics like the crude and RDS adjusted frequency, mean and standard deviation were calculated. Bivariate and multivariate logistic regression analyses were conducted to determine the strength of association between independent variables (risk factors) and the outcome variables (hepatitis B and C virus infection). Variable achieving a *P* value of <0.2 in the bivariate analysis were included in the multiple logistic regression model. Strength of association was measured using Adjusted Odd Ratios (AOR), precision of estimates determined using 95% confidence intervals, and a *P* value ≤ 0.05 was used as cut-off to determine statistical significance.

### Ethical Consideration

Ethical approval for the study protocol was obtained from the Scientific and Ethical Research Office (SERO) of the Ethiopian Public Health Institute (EPHI). Potential participants were told about the study purpose and procedures, potential risks, and protections using the local language. Written informed consent was obtained from each survey participant for the interview, blood sample collection, and storage of biospecimens for future testing.

## RESULTS

### Socio demographic characteristics

A total of 6085 FSW participated in the survey. Their median age [Interquartile Range (IQR = 8] was 25 years, and the highest number, 1980 (32.5%), were in age group 20-24 years and the lowest 615 (10.1%) in the age group 15-19 years (Table 1). A majority, 5031(82.7%), of them had formal education, 2946 (48.4%) were never married, and 4212 (69.2%) were pregnant a least once. A majority, 5694 (93%), of the respondents reported that selling sex was their main source of income, and 2066 (34%) of them had an average monthly income of Ethiopian Birr (ETB) 2500-4999 equivalent of U.S Dollar (USD) 60 – 120.

**Table 1:**
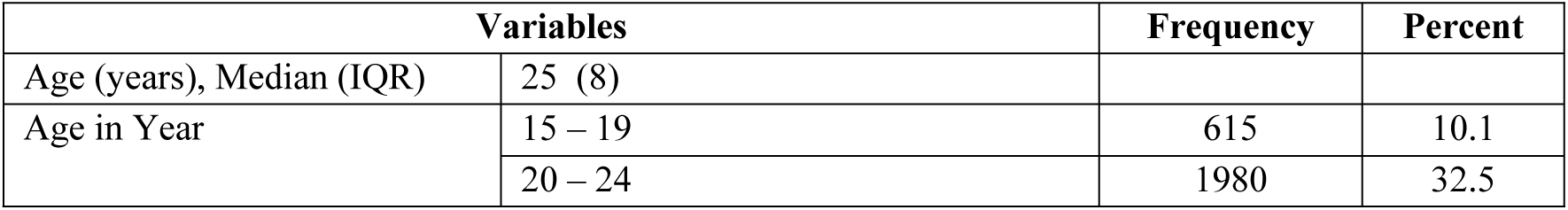
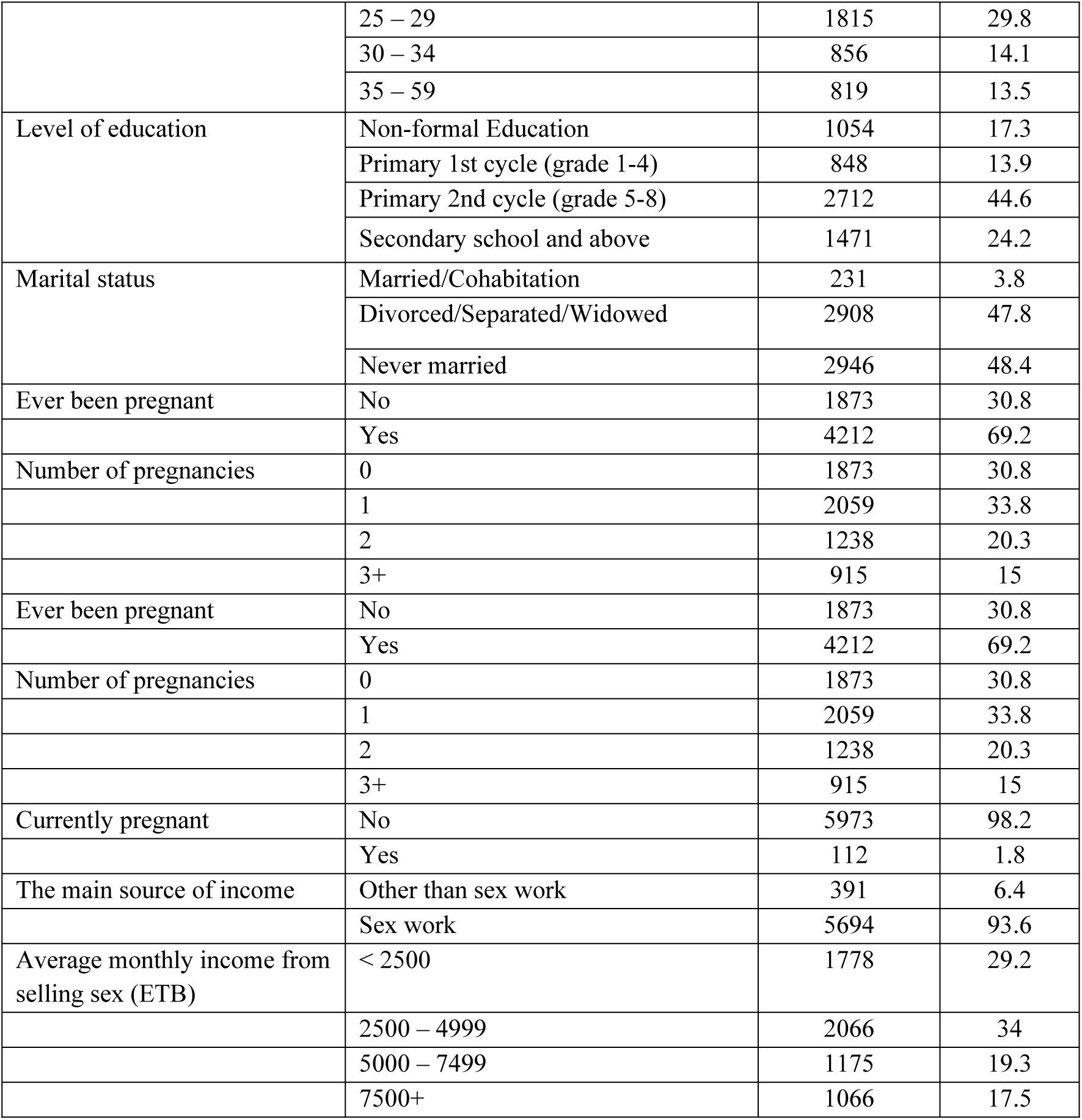
Socio-demographic characteristics among female sex workers in cities/towns, Ethiopia, 2020 (n= 6085)

### Distributions of FSW by study cities/towns

A majority, 1101 (18.11%), of the 6085 FSW resided in Addis Ababa, the national capital followed by Adama, 676 (11.1%). A smaller proportion of them were in other cities/towns (Table 2).

**Table 2:** Distributions of female sex workers by cities/town in Ethiopia, 2020 (n= 6085)

| Residence (City/Town) | Frequency | Percent |
| --- | --- | --- |
| Adama | 676 | 11.1 |
| Addis Ababa | 1101 | 18.1 |
| Hawassa | 522 | 8.6 |
| Gambella | 468 | 7.7 |
| Diredawa | 434 | 7.1 |
| Bahir Dar | 372 | 6.1 |
| Jimma | 254 | 4.2 |
| Mizan | 255 | 4.2 |
| Nekemite | 257 | 4.2 |
| Arba Minch | 251 | 4.1 |
| Combolcha/Dessie | 251 | 4.1 |
| Dilla | 251 | 4.1 |
| Diredawa | 434 | 7.1 |
| Gambella | 468 | 7.7 |
| Gonder | 250 | 4.1 |
| Logia/Semera | 251 | 4.1 |
| Shashemane | 250 | 4.1 |
| Harar | 242 | 4.0 |

### Sexual and behavioral characteristics

The median (IQR) age at first sex was 16 (3) years. The majority, 3384 (55.6%), of respondents had the first sex between the age of 16 and 20 years (Table 3). Some 2335 (38%) of the respondents had first sex with a person of their age, while 2425 (40%) of them had first sex with persons older by 5-10 years. The sexual practice was in hotels and bars in 2023 (33.2%) and was street-based in 1871 (30.7%). Alcohol dependence was report 2257 (37.2%) and chewing chat in 3827 (62.4%) of respondents during the last thirty days before the survey.

**Table 3:** Sexual and behavioral characteristics of female sex workers in cities /towns, Ethiopia, 2020 (n= 6085)

| Variables |  | Frequency | Percent |
| --- | --- | --- | --- |
| Age at first sexual intercourse (years), median (IQR) | 16 (3) |  |  |
| Age at first sexual intercourse (years), n (%) | ≤15 | 2430 | 39.9 |
|  | 16-20 | 3384 | 55.6 |
|  | ≥21 | 271 | 4.5 |
| Age at first sex selling | < 20 | 2328 | 38.3 |
|  | 20-24 | 2348 | 38.6 |
|  | ≥25 | 1406 | 23.1 |
| Age of first sex partner | 5 or more years younger | 73 | 1 |
|  | About the same age | 2335 | 38.4 |
|  | 5-10 years older | 2425 | 39.9 |
|  | More than 10 years older | 1252 | 20.6 |
| Location of sexual practice | Hotel/bar-based | 2023 | 33.2 |
|  | Street-based | 1871 | 30.7 |
|  | Home-based | 348 | 5.7 |
|  | Any type** | 661 | 10.8 |
| First sex experience | Wanted | 4738 | 77.9 |
|  | Forced | 1347 | 22.1 |
| Number of paying partners in the last 6 months, n (%) | ≤ 30 | 2308 | 37.9 |
|  | 31 – 60 | 1436 | 23.6 |
|  | 61 – 90 | 696 | 11.4 |
|  | ≥91 | 1645 | 27 |
| Ever had Anal intercourse, n (%) | Yes | 426 | 7 |
|  | Never | 5659 | 93 |
| Used condom during anal sex (n=426) | Yes | 252 | 59.2 |
|  | No | 174 | 40.8 |
| Consistent Condom utilization during the last 30 days with paying clients | Yes | 5119 | 84.1 |
|  | No | 966 | 15.9 |
| HBV among non-condom users during the last 30 days with paying clients | HBsAg positive | 23 | 2.4 |
|  | HBsAg negative | 943 | 97.6 |
| HBV among condom users during the last 30 days with paying clients | HBsAg positive | 134 | 2.7 |
|  | HBsAg negative | 4651 | 97.3 |
| Breakage of condom during the last 30 days | No breakage | 4260 | 70 |
|  | Experienced breakage | 1825 | 30 |
| In the last 30 days, used cigarettes or cigars | No | 5343 | 87.8 |
|  | Yes | 742 | 12.2 |
| Alcohol consumption level (AUDIT scores) | Not Risky | 2594 | 42.8 |
|  | Harmful, hazardous drinking | 1210 | 20 |
|  | Alcohol dependence indication | 2257 | 37.2 |
| Chewing khat in the last 30 days | Yes | 3827 | 62.9 |
|  | No | 2258 | 37.1 |
| Any drug used other than alcohol and khat in the last 30 days | Yes | 700 | 11.5 |
|  | No | 5382 | 88.4 |
| Used shisha in the last 30 days | Yes | 856 | 14.1 |
|  | No |  | 85.9 |

\*Alcohol Use Disorders Identification Test (long version)
\*\*Any type: Restaurant/cafe /cake bet, Local drink house (arake bet, tella bet, tej bet),
SPA/massage/beauty, Redlight

Condom use was practiced in 5119 (84%) FSW. Of the remaining 966 (16%) who were not using condom, 23 (2.4%) were HBsAg positive, and among those who reported using condoms systematically at every sexual intercourse, 134 (2.7) were HBsAg positive. HCVAb was positive in 4 and 23 of those who used condom consistently and inconsistently, respectively.

The first sex experience of the FSW was in 1347 (22.1%) forced, whereas in 4738 (77.9%) not forced. In the last six months preceding the study, 1645 (27%) of the FSW reported they had more than 90 clients. The majority, 5119 (84.1%), of the FSW utilized a condom during sex, and condom breakage was experienced by 1825 (30%).

### Prevalence of co-infection of HBV, HCV, syphilis and HIV among FSW

Of 1,140 FSW infected and 4945 not infected with HIV, 40 (3.5%) and of 117 (2.2%) were coinfected with HBV, respectively (Table 4). HCV coinfection was documented in 12 (1.1%) of those infected and 15 (0.3%) in those not infected with HIV. Syphilis was diagnosed in 339 (3.6%) of the FSW, and HBV and HCV coinfections with syphilis were seen in 3.8% and 2.1% among those infected and not infected with syphilis, respectively.

**Table 4:** Weighted prevalence of hepatitis B and hepatitis C co-infection with syphilis and HIV among female sex workers, Ethiopia, 2020 (N = 6085)

| Variables |  | Frequency | Percent |
| --- | --- | --- | --- |
| Depression level | Not depressed | 2468 | 40.6 |
|  | Mild depression | 2525 | 41.5 |
|  | Moderate to severe depression | 1092 | 17.9 |
| History of STI infection | Syphilis | 339 | 5.6 |
|  | Abnormal vaginal discharge | 887 | 14.6 |
|  | Syphilis HIV co-infection | 161 | 2.6 |
|  | Genital ulcer | 381 | 6.3 |
| HIV test result | Tested negative | 4945 | 81.3 |
|  | Tested new positive | 565 | 9.3 |
|  | Already known positive | 575 | 9.4 |
| Hepatitis B/HIV co-infection | HIV negative | 117 | 2.2 |
|  | HIV positive | 40 | 3.5 |
| Hepatitis C/HIV co-infection | HIV negative | 15 | 0.3 |
|  | HIV positive | 12 | 1.1 |
| Syphilis/HBV and HCV co-infection | HBV co-infection | 13 | 3.8 |
|  | HCV co-infection | 7 | 2.1 |

#### Prevalence of HBV and HCV

Of the 6085 FSW tested for HBV, 157 were tested positive for the HBsAg, a prevalence of 2.6% (95% CI (2.2, 2.8), and 27 FSW were positive for HCVAb, a prevalence of 0.5% (95% CI (0.4, 0.7). Only 5 (0.1%) of the participants were co-infected with both HBsAg and HCVAb. Among the 6085 participants in the study, a total of 184 (3%) were infected by HBV and HCV (Table 5).

**Table 5:** Weighted Prevalence of hepatitis B and hepatitis C infections among female sex workers, Ethiopia, 2020 (N = 6085)

| Characteristic | Overall (N=6085) | Percent (%) | 95% CI |
| --- | --- | --- | --- |
| HBV | 157 | 2.6 | 2.2-2.8 |
| HCV | 27 | 0.5 | 0.4-0.7 |
| HBV/HCV | 5 | 0.09 | 0.03-0.29 |
| HBV among non-condom users during sex (n=966) | 23 | 2.4 | 2.0-3.4 |
| HBV among condom users during sex (n=5119) | 134 | 2.6 | 2.2-2.8) |
| HCV among non-condom users during sex (n=966) | 4 | 0.4 | 0.2-0.8 |
| HCV among condom users during sex (n=5119) | 23 | 0.5 | 0.4-0.7 |

#### Factors associated with hepatitis B and C

Results of bivariate and multivariate logistic regression analyses are presented in Table 6. In the bivariate analysis, FSW with HBV infection had a significantly higher odds of being in the age groups 25-29 and 30-34, [COR=1.2; 95% CI (1.01, 3.93), *P*=0.045)] and [COR=2.35; 95% CI (1.14, 4.81), *P*=0.02)], respectively, compared with the age group 15-19 years. The odds of having over 90 sexual partners compared with those having under 30 partners in the past six months of being in age groups 20-24 or 25 years and above at first sex selling compared with those under 20, and of being HIV positive compared with being HIV negative was significant among FSW with HBV infection, [COR=1.6; 95% CI (1.07, 2.4), *P*=0.072)], [COR=1.68; 95% CI (1.15, 2.46), *P*=0.008)], [COR=1.64; 95%CI (1.06, 2.52), *P*=).025)] and [COR=1.72; 95% CI (1.09, 2.71), *P*=0.020)] respectively. In the multivariate logistic regression analysis, having more than 90 sexual partners in the past six months compared with those having less than 30 partners, being in the age groups 20-24 and ≥25 at first sex selling compared with those under 20 years, and being HIV positive, [AOR=1.66; 95% CI (1.11, 2.49), *P*= 0.013)], [AOR=1.67; 95% CI (1.14, 2.44), *P*=0.009)], [AOR=1.56; 95% CI (1.004 2.43), *P*=0.048)] and [AOR=1.64; 95% CI (1.03, 2.62), *P*=0.036)], respectively, are significantly and independently associated with HBV infection among FSW.

**Table 6:** Factors associated with hepatitis B and hepatitis C among female sex workers, Ethiopia, 2020

| Variable | Frequency | HBV (n=157 ) |  |  |  | HCV (n=27 ) |  |  |  |
| --- | --- | --- | --- | --- | --- | --- | --- | --- | --- |
|  |  | COR (95%C.I) | P-value | AOR (95%CI) | P-Value | COR (95%C.I) | P-value | AOR (95%CI) | P-value |
| Age | 15 - 19 | 1* |  |  |  |  |  |  |  |
|  | 20 - 24 | 1.15<br>(0.57, 2.33) | 0.390 |  |  |  |  |  |  |
|  | 25 - 29 | 1.2<br>(1.01, 3.93)* | 0.045 |  |  |  |  |  |  |
|  | 30 - 34 | 2.35<br>(1.14, 4.81)* | 0.02 |  |  |  |  |  |  |
|  | 35 - 59 | 1.51<br>(0.7, 3.26) | 0.3 |  |  |  |  |  |  |
| Number of sexual partner in the past 6 months | < 30 | 1* |  |  |  |  |  |  |  |
|  | 31 - 60 | 1.3<br>(0.84, 2.02) | 0.240 | 1.31<br>(0.84, 2.03) | 0.228 |  |  |  |  |
|  | 61 - 90 | **1.6**<br>**(0.96, 2.69)\*** | **0.072** | **1.66**<br>**(0.99, 2.79)\*\*** | **0.054** |  |  |  |  |
|  | 91+ | **1.6**<br>**(1.07, 2.40)\*** | **0.021** | **1.66**<br>**(1.11, 2.49)\*\*** | **0.013** |  |  |  |  |
| Average monthly income from selling sex in ETB | < 2500 | 1* |  |  |  |  |  |  |  |
|  | 2500 – 4999 | 1.16<br>(0.76, 1.77) | 0.496 |  |  |  |  |  |  |
|  | 5000 – 7499 | 1.49<br>(0.94, 2.36) | 0.089 |  |  |  |  |  |  |
|  | 7500+ | 1.37<br>(0.85, 2.22) | 0.198 |  |  |  |  |  |  |
| Moderate to severe depression | Not depressed | 1* |  |  |  |  |  |  |  |
|  | Mild Depression | 0.72<br>(0.51, 1.02)* | 0.067 |  |  | 0.98<br>(0.39,2.47) | 0.961 |  |  |
|  | Moderate to severe depression | 0.85<br>(0.55, 1.32)* | 0.474 |  |  | 2.27<br>(0.90-5.74) | 0.083 |  |  |
| Number of cities worked sex selling in the last three years | Same town |  |  |  |  |  |  |  |  |
|  | 1 more town | 1.48<br>(0.97, 2.27) | 0.069 |  |  |  |  |  |  |
|  | 2 or more towns | 1.48<br>(0.83, 2.66) | 0.186 |  |  |  |  |  |  |
| Number of non-paying partners in the past 6 month | Never |  |  |  |  |  |  |  |  |
|  | Only one | 2.34<br>(0.86, 6.38) | 0.096 |  |  |  |  |  |  |
|  | 2 and more | 1.98<br>(0.70, 5.64) | 0.198 |  |  |  |  |  |  |
| Age at first sex | 15 or less |  |  |  |  | 0.24<br>(0.08,0.70) | 0.009 | **0.21**<br>**(0.07,0.61)\*\*** | **0.005** |
|  | 16 – 20 |  |  |  |  | 0.17<br>(0.06,0.50) | 0.001 | **0.18**<br>**(0.061,0.53)\*\*** | **0.002** |
|  | 21+ |  |  |  |  | 1* |  |  |  |
| Age at first sex selling | < 20 | 1* |  |  |  |  |  |  |  |
|  | 20 – 24 | **1.68**<br>**(1.15, 2.46)** | **0.008** | **1.67**<br>**(1.14, 2.44)\*\*** | **0.009** | 0.59<br>(0.22,1.64) | 0.314 |  |  |
|  | 25+ | **1.64**<br>**(1.06, 2.52)** | **0.025** | **1.56**<br>**1.004, 2.43)\*\*** | **0.048** | 1.83<br>(0.77,4.31) | 0.169 |  |  |
| HIV Test Result | Tested Negative | 1* |  |  |  |  |  |  |  |
|  | Tested New Positive | 1.28<br>(0.76, 1.14) | 0.349 | 1.249<br>(0.74, 2.09) | 0.410 | 2.93<br>(1.06-8.10) | 0.038 | 2.05<br>(0.71,5.92) | 0.186 |
|  | Known Positive | **1.72**<br>**(1.09, 2.71)\*** | **0.020** | **1.64**<br>**(1.03, 2.62)\*\*** | **0.036** | **4.05**<br>**(1.64-9.98)\*** | **0.002** | **2.85**<br>**(1.10,7.37)\*\*** | **0.031** |
| Syphilis | Non-Reactive | 1* |  |  |  |  |  |  |  |
|  | Reactive | 1.55<br>(0.87, 2.77) | 0.137 |  |  | **6.03**<br>**(2.53-14.38)** | **0.000** | **4.38**<br>**(1.73,11.11)** | **0.002** |

1\*- reference ( used as constant); \*- significant on Bivariate analysis; \*\*- significant at multivariate analysis

In the bivariate analysis, FSW with HCV infection had a lower odds of being in the younger aged group of 15 years or under was significantly higher [COR=0.24, 95% CI (0.08,0.70), *P*= 0.009)] and 16-20 years [COR=0.17; 95% CI (0.06,0.50), *P*= 0.001)] compared those aged above 20 years. The odds of being newly HIV positive or known HIV positive compared with being HIV negative and being positive for syphilis compared with being negative for syphilis was significant [COR=2.93; 95% CI (1.06-8.10), *P*= 0.038)], [COR=4.05; 95% CI (1.64-9.98); *P*=0.002)] and [COR=6.03; 95% CI (2.53, 14.38), *P*= 0.00)], respectively. In the multivariate logistic regression analysis, FSW with HCV infection is significantly and independently associated with age at first sex of 15 years or less [AOR=0.21; 95% CI (0.07, 0.61), *P* = 0.005)] and age 16-20 years, compared with age of 25 years and above [AOR=0.18; (0.061, 0.53), *P*= 0.002)] known HIV status [AOR=2.85; 95% CI (1.10, 7.37), *P* = 0.031)] and being positive for syphilis [AOR=4.38; 95% CI (1.73,11.11), *P* = 0.002)]

## DISCUSSION

Of the 6085 FSW enrolled in our study in 2020, the prevalence of HBV among FSW was 2.6% and of HCV was 0.5%, which are comparable with a similar, previous study. Based on the WHO HBV infection prevalence classification - high (>8%), intermediate (2-8%), and low (<2%) (24), HBV infection prevalence in Ethiopia is the intermediate category. This translates to almost one in every thirty-seven FSW having HBV infection. The prevalence we have identified in the Ethiopian setting is lower than the prevalence reported from the rest of Africa and South-East Asia (5%), but higher than the prevalence in the Americas and Eastern Mediterranean (1%). This contributes to the 1.5 million new HBV infections occurring globally each year and will have major implications to the epidemiology of chroming HBV infection; nearly 3.6% of the world’s population (257-296 million) have chronic HBV infection, a chronic HBV infection prevalence of 0.01-2% in the United Kingdom, United States, Canada, Western Europe, and Japan, and over 8% in most SSA and Western Pacific regions (25-28).

In countries with high and middle-income, HBV transmission is more perinatal and horizontal, whereas, in low-income countries, the transmission occurs through drug injection and high-risk sexual behaviors (29). Africa and Asia have the highest endemicity of HBV, though effective vaccination programs have pushed the burden towards moderate or low endemicity Most countries in Africa have high endemicity, with the exception of Tunisia and Morocco, which have moderate endemicity (30,31). The WHO global hepatitis strategy aims to reduce new hepatitis infections by 90% and deaths by 65% between 2016 and 2030 (32), and according to WHO, 80% of people with hepatitis live without prevention, testing, and treatment of HBV infection (33).

There is much variation in the prevalence HBV infection across countries, including those which have a lower prevalence than ours like Mexico 0.2% (34), Iran 1.1% (35), Greece 1.3% (36), Brazil 0.7% (37), those with similar prevalence to ours like Rwanda 2.5% (38), and countries in SSA having much higher prevalence than ours like Nigeria 17.1% (39), Kenya, 13.3% (40), and Ghana 15.0% (41). Similarly, isolated and limited studies on HBV conducted in different cities/towns in Ethiopia at various times reported varying prevalence by site and year of study - Dessie 13.1% (11), Hawassa 9.2% (14), Gonder 28.9% (15), and Mekelle 6% (16). This variation could largely be explained by the differences in sociodemographic characteristics of study populations, study settings, sample size, and sampling methods focusing on high-risk population groups.

Our study showed that HBV prevalence was significantly associated with the age groups 25-29 years and 30-34 years compared to the age group 15-19 years in the bivariate analysis, but these did not achieve significant independent association in the multivariate analysis (P > 0.05). This is consistence with the finding by Forbi JC. et al (39) who reported that, although the prevalence of HBV is highest among FSW in the 30-35 years age group, it is not statistically significant. In contrast to this finding Vázquez-Martínez, et al (42). Recognized that being above 30 years of age is significantly associated with HBV infection. the duration of staying as CSW among FSW increased the infection risk of HBV with increased age. In Our study only individuals above 15-years were recruited. the finding implies awareness creation and high engagement of health workers will be needed to vaccinate high-risk groups like FSW.

Having more than 90 sexual partners in the past six months on a monthly average of 15, compared with those having less than 30 partners in the past six month, almost one partner on monthly average was independently associated with HBV infection. The positive association of HBV exposure with having a greater number of clients indicates that FSW were vulnerable to infection through sexual risks. This is not surprising as sexual transmission is an important route of transmission of HBV infection. Generally speaking, the reported average number of clients who had a history of multiple sex partners may increase the probability of having sex with an infected partner during the acute phase of infection this study was in agreement with study conducted by the WHO 2014 Global Network of Sex Work Projects, Prevention and treatment of HIV and other STS for sex workers in low and middle-income countries and study result from Ethiopia, India, Brazil, Egypt and Japan (43-49).

Being in the age groups 20-24 and 25 years and above at first sex selling was significantly and independently associated with HBV infection compared with those under 20 years. This finding was inconsistency with study reports from Nigeria and Mexico, which support early age of sexual activities increases the risk of HBV infection (39,42). This variation might be an indication for FSW in this study had sexual involvement at an early stage and did not get HBV vaccination during their childhood because the national viral hepatitis prevention and control program was initiated in 2016 in Ethiopia. Thus, most of our study participants did not have the opportunity for early vaccination. However, in the previous studies conducted in Nigeria and Mexico, HBV vaccination was given in early childhood. In addition, the females in Ethiopia are economically independent of their families at the age of 20 years or above. In order to increase their daily expenses the FSW might be sexually active and engaged with many partners which increases their exposure to HBV. We could not find any report that indicated the significant association of being HIV positive with HBV infection alone. However, the study reported from Rwanda on Syphilis and HIV prevalence and associated factors to their co-infection among FSW indicated that HBV infection is an independent predictor of HIV and syphilis co-infection, and the odd of having an HBsAg-positive test is 2.09 times higher in syphilis/HIV co-infection FSW(38). This result was almost similar to our finding in which Being HIV positive compared with being HIV negative was significant among FSW was independently associated with HBV infection in which being HIV positive was 1.64 times more likely to be infected by HBV than FSW who were HIV negative. The presence of HBV infection among FSW might be due to a higher risk of developing hepatotoxicity following the initiation of antiretroviral therapy or a lower CD4 T-cell count.

We found that the overall prevalence of HCV among FSW was 0.5%, a finding similar to that reported form Nairobi, Kenya (0.76%) (43), and a previous study in Ethiopia (0.7%) (50). Our finding is also concurs with the prevalence ranging between 0-1.4% reported from the united States and Europe (51,52) as well as the global average prevalence of HCV infection (0.8%) (53). In contrast our finding is lower than the prevalence reported from Iran of 6.2 % (54), Ghana of 2.8% (55), and Port Harcourt, Nigeria 2.9% (56). Similarly, syphilis seroprevalence of 5.6% from this study is higher than that of Ghana 7.5% (57), Tanzania 12.7% (58), and a previous report from Ethiopia 1.3% (50). Our finding indicates that the prevalence of HCV was relatively lower compared to similar finding reported elsewhere. The reason for the lower report might be the improvement in technology where the current screening reagent to be more specific and reliable, and could also be an indication for the improvement of the national prevention or a treatment programs for HCV. It could also be an indicator that there are geographical discrepancies in prevalence of HCV infection.

Our finding indicates, that FSW with HCV infection had a lower odds of being in the younger aged group of 20 years or under is significantly and independently higher compared to those aged above 20 years. The rise in anti-HCV positivity with age in this group might be the result of continuous exposure to the virus. Our finding suggests that HCV seroprevalence is lower among the younger FSW and this might be a reflection of lower exposure to sexual risk behaviors and the small sample size in the younger age group which need be taken into consideration, Concurring with our finding, a previous study also indicated that the low HCV seroprevalence in younger CSW (59).

Mainly because of the nature of HIV route of transmission, many HBV or HCV-infected individuals were co-infected with HIV. Our study adds to the evidence showing that co-infection of HIV and viral hepatitis occurs frequently largely because of their mode of transmission. The odds of being newly diagnosed with HIV positive or having a known HIV positive status among FSW with HCV infection was significantly and independently higher compared with HIV negative FSW with HCV. FSW who were known HIV positive were 2.85 times at higher risk of being HCV infected compared to those who were HIV negative. This finding was supported with a study conducted in Burkina Faso, West Africa, and the results of a systematic review on HIV and the hepatitis virus co-infection in studies from SSA, which reported that those who were HIV infected were estimated to be at higher risk of HCV coinfection compared to HIV seronegative people (AOR = 5.59) and (RR=1.60), respectively (17,60). The hepatitis C virus is usually spread when someone comes into contact with blood from an infected person. This can happen through sharing drug injection equipment, though drug injection is uncommon in Ethiopia (61). Our finding suggest that the route of transmission for HCV among sexually active individuals is commonly sexual. After stratification by HIV status, HCV prevalence among women of the general population was identical to that of FSW, suggesting that HCV sexual transmission is not common in this population and that HIV infection does not enhance susceptibility to HCV sexual transmission (62). However, the significant association between HCV and HIV infection among FSW has not been clearly described in published studies and this needs further investigation.

The odds of being positive for syphilis compared with being negative was significantly and independently associated with being HCV positive among FSW. In agreement with our finding, a previous study also indicated that HCV sero-reactivity or positivity was significantly predicted and associated with syphilis sero-reactivity or positivity (63). Moreover, a study conducted by Tessema B, et al (50) suggested that the highest rate of co-infection and the statistically significant relationship between HCV and syphilis infections might be due to the fact that these pathogens share common modes of transmission and risk groups. These findings are in line with our finding showing that FSW with HCV infection were 4.4 times more likely to be syphilis sero-reactive or positive than those who were syphilis non-reactive or negative. As HCV positive FSW were at a higher risk for having syphilis, prevention mechanisms and intervention need to be instituted among FSW to decrease further transmission of HCV and syphilis to the general population.

Of 6085 participants included in our study,184 (3%) had results for both HBsAg and HCVAb. The prevalence of HBV/HCV, HIV/HBV, and HIV/HCV co-infection among HIV positive FSW in the present study was 3.6%, 1.3%, and 0.09%, respectively. A similar finding was reported from the Global Prevalence of HBsAg and HIV and HCV Antibody study, which showed that the prevalence of HIV/HBV and HIV/HCV co-infections among FSW was 3% and 1%, respectively (31).

The previous prevalence estimates of HBV among general population in Ethiopia ranged from 8%-12%, and HCV prevalence estimated at greater than 2.5% (13). These findings were higher than our finding among FSW, a group at a much high risk of getting HBV/HCV and STI. It appears that the national estimates of these infections among the general population could have been overestimated. Overall, the different studies conducted in Ethiopia on HBV and HCV have produced varying seroprevalence estimates. The studies have been conducted in different population groups carrying varying risks, utilized different sample sizes, and using different laboratory screening methods, some with, and others without laboratory confirmatory testing to arrive at seroprevalence estimates. In addition, the studies were conducted in different geographic settings. These pose a major challenge in reaching a consensus on the prevalence of hepatitis viruses in Ethiopia. In light of these limitations, a large-scale seroprevalence and epidemiological studies need to be done to ensure a more robust and current national seroprevalence estimate.

The main strength of our study was the inclusion of participants from high-risk groups and being the first report on the prevalence of viral hepatitis B and C among FSW in Ethiopia at national and regional levels. The result from this study are among the first in the country and in the region, to explore both hepatitis B and C among FSW, to categorize high-risk and vulnerable population, in order to fill the gap on data of virial hepatitis in Ethiopia among key populations and in addition, this study was conducted in a large sample of FSW across 16 cities/towns of the country, which makes the findings generalizable to FSW of Ethiopia.

Nevertheless, this study also had limitations. First, as part of the national surveillance, the survey done in this round had targeted only provincial (regional) capitals, and major towns. Even in the selected cities, the presence of harder-to-reach sex worker groups like home-based sex workers might not be fully accounted for lack of detailed city maps for all regional capitals, and street names were challenges in the mapping and presentation of the results from the size estimation study. Second, as most studies were set in urban areas and as FSW were predominated included in the study, the generalizability of the results need to be considered with some caution.

## Conclusion

This study reveals an intermediate prevalence of HBV and a low prevalence of HCV infections among FSW in cities and town in Ethiopia. Our prevalence finding is lower than the estimate among the general population that was reported by previous studies. While this might have been influenced by difference in methodologies, our finding may also suggest that the coverage of hepatitis related interventions among FSW has been effective, but require further strengthening to control HBV and HCV transmission among FSW. Scaling-up interventions for FSW such as HBV vaccination, reducing number of sexual partners, increasing condom distribution and regularly monitoring and screening for these infections and other STI among FSW is needed. FSW who are HBV and HCV infected should be informed about the transmission routes and methods to prevent further spread of the viruses. FSW testing HBV negative but not yet got vaccinated need to receive the vaccine since they were at high-risk of contracting the infection. Further epidemiological studies to determine the prevalence and determinants of HBV, HCV and other STI among different population groups are suggested.

## Data Availability

By following the Ethiopian Public Health Institute's data usage laws and regulations, data-set documentation is available for immediate download and datasets are available upon request for access and depository

## ACKNOWLEDGMENT

The authors would like to acknowledge the Ethiopian Public Health Institute for providing materials support during this project implementation. We also thank Ethiopian Public Health institute, National HIV/AIDS surveillance and laboratory treatment center staff members for their cooperation during data collection process. We would also like to acknowledge all the Female sex workers and their partners whose data were used in this study, and all the healthcare workers who took part in the educating and treating of the included Female sex workers.

## AUTHORS CONTRIBUTION

**Conceptualization**: Birra Bejiga Bedassa

**Data curation**: Birra Bejiga Bedassa, Gemechu Gudeta Ebo, Feyiso Bati Wariso, Jemal Ayalew, Saro Abdella, Sileshi Lulseged

**Formal analysis**: Birra Bejiga Bedassa, Gemechu Gudeta Ebo, Jemal Ayalew.

**Funding acquisition**: Saro Abdella, Getachew Tollera, Tsigereda Kifle

**Methodology**: Birra Bejiga Bedassa.

**Project administration**: Jaleta Bulti Tura. Getachew Tollera, Tsigereda Kifle, Saro Abdella

**Resources**: Jaleta Bulti Tura, Saro Abdella

**Software:** Jemal Ayalew, Gemechu Gudeta Ebo, Feyiso Bati Wariso

**Supervision**: Birra Bejiga Bedassa, Gemechu Gudeta Ebo, Jaleta Bulti Tura

**Validation:** Birra Bejiga Bedassa, Sileshi Lulseged

**Visualization**: Birra Bejiga Bedassa, Sileshi Lulseged.

**Writing – original draft**: Birra Bejiga Bedassa, Gemechu Gudeta Ebo, Feyiso Bati Wariso

**Writing – review & editing**: Gemechu Gudeta Ebo, Jemal Ayalew, Saro Abdella, Sileshi Lulseged

